# Elevated Levels of IL-9 Fail to Suppress Pathogenic T helper 17 cells in Sjögren’s Disease

**DOI:** 10.64898/2025.12.19.25335657

**Authors:** Alexandria Voigt, Yiran Shen, Patricia Glenton, Astrid Rasmussen, Robert H. Scofield, Kiely Grundahl, Christopher J. Lessard, A. Darise Farris, Cuong Nguyen

**Affiliations:** Department of Infectious Diseases and Immunology, College of Veterinary Medicine, Gainesville, FL; Arthritis and Clinical Immunology Program, Oklahoma Medical Research Foundation, Oklahoma City, OK; Department of Oral Biology, College of Dentistry; Center of Orphaned Autoimmune Diseases, University of Florida, Gainesville, FL, USA

**Keywords:** Interleukin 9, Sjögren’s Disease, Regulatory T cells, T helper 17 cells

## Abstract

Sjögren’s disease (SjD) is a chronic autoimmune disorder characterized by inflammation of the exocrine glands, leading to dry mouth and dry eyes. This study investigates the role of interleukin-9 (IL-9) and T helper 9 (Th9) cells in the pathogenesis of SjD. We found that serum IL-9 levels were significantly elevated in SjD patients and correlated with clinical laboratory parameters, including autoantibody production. In a mouse model of SjD, IL-9 and Th9-associated cytokines were also elevated, and Th9 cells were enriched in the salivary glands. Our results suggest that IL-9 is produced by multiple cell types, including macrophages, CD4^+^ T cells, and NK cells, and that Th9 cells contribute to the development of SjD by promoting inflammation and autoantibody production. We also found that Th9 and Th17 polarization conditions increased Th2 and Th17 cells in SjD mice, indicating a shared epigenetic program that renders T cells permissive to multiple differentiation pathways. Anti-IL-9 treatment had a sex-dependent effect, reducing autoantibody production in male mice but worsening focal glandular infiltration in female mice. Our findings suggest that IL-9 plays a complex role in SjD pathobiology, contributing to both local immunoregulation and systemic autoantibody response. Overall, this study offers new insights into the role of IL-9 and Th9 cells in SjD, highlighting the potential for therapeutic targeting of the IL-9/Th9 axis in the treatment of this disease.

## INTRODUCTION

Sjögren’s Disease (SjD) is a chronic autoimmune disease characterized by inflammation of the exocrine glands, primarily the salivary and lacrimal glands, leading to dry mouth (xerostomia) and dry eyes (xerophthalmia). The disease can also manifest with various extraglandular symptoms, including fatigue, arthralgias, and an increased risk of lymphoma. SjD is a complex disorder, and its pathogenesis involves the interplay of multiple immune cells, cytokines, and other molecules. Previous studies have supported the pathogenic roles of effector T cells, such as T helper 1 (Th1), Th2, and Th17 cells in SjD. One of the other key players in the immune system is the Th9 cell, a subset of CD4+ T cells that produces interleukin-9 (IL-9). Th9 cells have been implicated in the pathogenesis of several autoimmune diseases, including SjD.

IL-9 is a pleiotropic cytokine that plays a crucial role in regulating immune responses, including the activation and differentiation of various immune cells. It is involved in various immune responses, including autoimmunity, where it promotes the activation and differentiation of autoreactive T cells, leading to the production of autoantibodies and tissue damage. Th9 cells also contribute to inflammation by recruiting immune cells to the site of infection or injury and enhancing the production of other pro-inflammatory cytokines. Additionally, they play a role in antitumor immunity by promoting the activation and expansion of tumor-specific T cells. Additionally, Th9 cells are implicated in parasite immunity, tissue repair, and immune regulation, producing anti-inflammatory cytokines like IL-10 to suppress immune cell activation and promote tolerance. The development and function of Th9 cells are regulated by transcription factors such as PU.1, IRF4, and BATF, which control the expression of IL-9 and other genes involved in Th9 cell development and function, ultimately contributing to a complex role in the immune response.

IL-9 and Th9 cells have been implicated in the pathogenesis of several other autoimmune diseases, including rheumatoid arthritis (RA), systemic lupus erythematosus (SLE), and multiple sclerosis (MS). Studies have found that Th9 cells and IL-9 are upregulated in the peripheral blood and synovium of patients with RA^1^, and are associated with the severity of the disease and biomarkers, such as the Disease Activity Score^2^. Similarly, high expression of IL-9 levels and the percentages of CD4^+^IL-9^+^ T cells correlate with disease activity and severity (SLEDAI)^3^. A recent study has shown that PU.1 transcription factor and IL-9 form a positive feedback loop in RA, in which PU.1 directly binds to the IL-9 promoter, activating IL-9 transcription, and Th9-derived IL-9 induces PU.1 via the IL-9R-JAK1/STAT3 pathway^4^. In the context of SjD, IL-9 and Th9 cells have been shown to play a significant role in the disease’s pathogenesis. Studies have demonstrated that IL-9 levels are elevated in the serum and salivary glands of patients with SjD. Furthermore, Th9 cells have been found to be enriched in the salivary glands of SjD patients, which can be speculated to lead to the recruitment of immune cells to the salivary glands and the development of inflammation. However, the mechanisms by which IL-9 and Th9 cells contribute to the development of SjD are not fully understood. In this study, we aimed to determine the clinical association of SjD and IL-9. Using animal models, we examined the cellular source of IL-9 in the salivary glands and evaluated the immunological role of IL-9/Th9 in SjD.

## MATERIALS AND METHODS

### Human Subjects

The human subjects were participants in a large cohort of sicca patients evaluated at the Sjögren’s Research Clinic at the Oklahoma Medical Research Foundation (OMRF^5^. All the tests necessary for classification by AECG^6^ and ACR-EULAR^7^ criteria were performed in addition to the collection of detailed clinical and serological measures, as previously described^5^. The definition of the SjD case or control was based on the AECG classification criteria. Studies were approved by the Institutional Review Board of the Oklahoma Medical Research Foundation. The participants were provided written informed consent prior to entering the study. The study was conducted in accordance with current regulations that protect human subjects participating in research, including HIPAA and the Declaration of Helsinki.

### Mice

B6 and B6.NOD-*Aec1Aec2* (SjD) mice were bred and maintained under specific pathogen-free conditions in the animal facility of the University of Florida. All the procedures related to the use of animals were approved by the University of Florida Institutional Animal Care and Use Committee (IACUC) under the approved protocol (IACUC202300000418). All animals were maintained on a 12-hour light-dark schedule and provided food and acidified water *ad libitum*. Mice were anesthetized with isoflurane and euthanized by cervical dislocation; their organs and tissues were freshly harvested for analyses

### Cytokine Assays

The human serum cytokine levels were determined using a customized multiplex assay kit (BioRad, Hercules, CA) per the manufacturer’s instructions. In brief, sera samples were diluted 1:4 in the provided assay buffer. A 96-well assay plate was prewet with 100 μl of assay buffer. Pre-mixed beads were added to each well in a 50 μl volume and then washed, using a magnet for bead retention, before adding 50 μl of the diluted sample, standard, or blank. The assay plate was incubated for 30 min by shaking at room temperature in the dark. The plate was then washed before the addition of the detection antibody. Following the detection antibody, the plate was washed by adding 50 μl of Streptavidin-PE per well. The samples were incubated for 10 min by shaking. The plate was washed one final time, and 125 μL of assay buffer was added. The results were acquired using BioRad Magpix. The data were analyzed using the Bio-Plex Manager software (BioRad, Hercules, CA). The mouse serum cytokine measure was determined using a service provided by Olink (Olink, Uppsala, Sweden). The Olink Target 48 Mouse Exploratory was used to measure 48 circulating proteins from the eluates, which were assessed by proximity extension assays (PEA). Only the Th9-associated cytokines were presented in the study.

### Anti-IL-9 Antibody Treatment

Over the course of six weeks, B6.NOD-*Aec1Aec2* mice received an intraperitoneal (i.p.) injection of either an anti-IL-9 antibody (BE0181, BioXCell, Lebanon, NH) or an isotype antibody control (BE0085, BioXCell, Lebanon, NH) five times per week. During the initial two weeks, mice were injected with 40 ug per mouse, and during the final 4 weeks, the mice received a 100 ug dose.

### Saliva Flow Rate

Saliva flow rate (SFR) was recorded prior to beginning (baseline), then every two weeks, as previously described. Briefly, mice were weighed and given an intraperitoneal (i.p.) injection of 100 μl isopreterenol (0.2Lmg/1Lml of PBS) and pilocarpine (0.05Lmg/1Lml of PBS) to stimulate saliva production. After allowing the secretagogue to take effect for one minute, saliva was collected from the oral cavity with a pipette for ten minutes, with a one-minute break at the midpoint. The final SFR measurement was collected three days prior to euthanasia. Saliva was briefly centrifuged to allow for an accurate measurement of the volume, which was then divided by the weight of the individual mice and the length of time for collection, resulting in the units for SFR being reported as μL/g/10 minutes.

### Antinuclear Antibody Determination

Antinuclear antibodies were detected in the sera of mice per the manufacturer’s instructions (ImmunoConcepts, Sacramento, CA). Briefly, sera were diluted 1:40 in PBS and incubated on HEP-2 ANA slides for 30 minutes. After washing, the secondary antibody, goat anti-mouse IgG AF488 (Invitrogen, Waltham, MA), was incubated on the slide for 30 minutes. Afterwards, slides were sealed with Vectashield DAPI medium (Vector Laboratories, Burlingame, CA), and a glass coverslip was mounted. The ANA staining pattern was observed at 400x magnification using a Nikon Ti-E fluorescent microscope with an exposure time of 200 ms (Nikon, Tokyo, Japan).

### Histological Examination

At the endpoint, mice were anesthetized with isoflurane and euthanized by cervical dislocation. Salivary glands were isolated, placed in histology cassettes, and fixed in 10% phosphate-buffered formalin for 24 hours prior to outsourcing of processing at HistoTech (Gainesville, FL). There, tissues were paraffinμ-embedded, sections were cut at a 5 μm thickness, and mounted onto slides. Additionally, hematoxylin & eosin (H&E) staining was performed on these slides following deparaffinization via successive immersion in xylene and ethanol. H&E-stained sections were analyzed under a Nikon microscope at 200x magnification. Focal scores were determined by enumerating lymphocytic infiltrates of > 50 leukocytes for a single histological section per mouse.

### Flow cytometry

There were several different studies using an array of tissues, so firstly, here is how the tissues were processed. Briefly, salivary glands were dissociated using a MACS dissociator tube, then incubated in RMPI complete media (10 minutes, 37°C). Media containing single cells was poured into a fresh tube while the remaining tissue received fresh, warm media, and the above was repeated twice more. Both the single cell suspension and the remaining tissue were poured through a 70 μm cell strainer to remove debris. Similarly, draining cervical lymph nodes were mashed through a 70 um cell strainer using the plunger of a 3 mL syringe and washed. Spleens were mashed through a 70 um cell strainer using the plunger of a 3 mL syringe and washed.

Cells were resuspended in 7 mL of a % ammonium chloride solution and incubated (7 minutes), then centrifuged (300 x g, 10 minutes), followed by a wash. The globe of the eye was excised from the socket, and blood was collected into a heparin-treated tube. 5 mL cold RBC lysis buffer (0.802% NH4Cl, 0.084% NaHCO3, 0.037% EDTA) was incubated with 150 uL heparinized blood (13.5 min, room temperature). After washing twice with DPBS, cells were resuspended in 100 μL FACS buffer. After isolating these single cell suspensions, cells were resuspended in RMPI, stimulated with PMA, Ionomycin, and Golgi Stop, and incubated (4 hours, 37°C, 5% CO_2_). Cells were blocked with FC Block (Biolegend, San Diego, CA) per manufacturer’s instructions, washed, and incubated with 50 ng/mL PMA (Sigma-Aldrich, St. Louis, MO), 500 ng/mL Ionomycin (Sigma-Aldrich, St. Louis, MO), and Golgi Stop (BD Biosciences Franklin Lakes, NJ, Franklin Lakes, NJ) (3, hours, 37°C, 5% CO_2_).. For evaluation of source cells of IL-9, four panels were utilized for PerCP-eF710 anti-NKp46 (eBioscience 29A1.4, San Diego, CA), FITC anti-CD19 (Biolegend ID3/CD19, San Diego, CA), BV785 anti-CD11b (Biolegend M1/70, San Diego, CA), PE anti-CD11c (Biolegend HL3, San Diego, CA), PerCPP-Cy5.5 anti-CD4 (Biolegend GK1.5, San Diego, CA), FITC anti-CD8a (BD Biosciences 53.6-7, Franklin Lakes, NJ), PE anti-TCR gamma/delta (eBioscience, eBio GL3, San Diego, CA), APC anti-CD103 (Biolegend 2E7, San Diego, CA), BV605 anti-CD3e (Biolegend 145-2C11, San Diego, CA), BV785 anti-CD3e (Biolegend 145-2C11, San Diego, CA), APC anti-IFN_γ_ (Biolegend XMG1.2, San Diego, CA), AF488 anti-IL-17 (BD Biosciences TC11-18H10.1, Franklin Lakes, NJ), PerCP-eF710 anti-ROR_γ_T (eBioscience B2D, San Diego, CA), APC-Cy7 anti-CD25 (Biolegend PC61, San Diego, CA), AF647 anti-CD39 (Biolegend Duha59, San Diego, CA), PE-Cy5.5 anti-FoxP3 (eBioscience FJK-16s, San Diego, CA), PE anti-IL-10 (Biolegend JES5-16E3, San Diego, CA), and BV421 anti-IL-9 (Biolegend RM9A4, San Diego, CA). Flow cytometry was performed on BD Fortessa with analysis on FlowJo X. (FlowJo, Waltham, MA).

### Salivary Gland Single-Cell Analysis

The scRNA-seq dataset (GSE268532) was processed using the Seurat package (v5) in R. Filtered gene expression matrices from Cell Ranger Multi were imported, and Seurat objects were created for each sample. Quality control filtering was applied with the following thresholds: >200 and <6,000 detected genes per cell, <30,000 UMIs per cell, and <15% mitochondrial gene expression. Doublets were identified and removed using scDblFinder. Data integration across samples was performed using Harmony to correct for batch effects between samples and sex. Principal component analysis was conducted, and the optimal number of dimensions was determined using elbow plots. Clustering resolution was optimized using clustree visualization, followed by Uniform Manifold Approximation and Projection (UMAP) for dimensionality reduction and visualization. Cell type annotation was performed based on the expression of canonical marker genes. Gene expression percentages and average expression levels were calculated for each cell type-phenotype combination using the AverageExpression function. UCell scores for Th9 transcriptional signatures were calculated using genes from previously published Th9 transcription factor networks^8,9^, employing Mann-Whitney U statistical analysis to generate individual cell scores for signature enrichment.

### Treg suppression assay

To evaluate the relationship between Treg and T helper cells in relation to IL-9 or anti-IL-9 regulation, B6 and B6.NOD-*Aec1Aec2* mice were anesthetized with isoflurane and euthanized by cervical dislocation; spleens were processed as described above to achieve the single cell suspension. Splenocytes were then processed with MACS columns on the MACS magnet to isolate antigen-presenting cells (APCs), Tregs, or naive T cells. Splenocytes were labeled with the following: CD8 (Biolegend 5H10, San Diego, CA) CD45 (BD Biosciences RA3-6B2, Franklin Lakes, NJ), CD11b (Biolegend M1/70, San Diego, CA), CD11c (BD Pharmingen HL3, Franklin Lakes, NJ), TCR gamma/delta (Biolegend GL3, San Diego, CA), Ter-119 (Biolegend TER-119, San Diego, CA), and CD49 (Biolegend DX5, San Diego, CA)After labeling with anti-PE microbeads, a LD column (Miltenyi Biotec, Bergisch Gladbach, Germany) was utilized to deplete these populations and flow-through was collected. Isolated cells were labeled with PE anti-CD25 (Biolegend 3C7, San Diego, CA) followed by anti-PE microbeads (Miltenyi Biotec, Bergisch Gladbach, Germany), and an MS column (Miltenyi Biotec, Bergisch Gladbach, Germany) was utilized to isolate the Tregs. The flow-through was also collected and stained with anti-CD62L microbeads, then loaded onto the column for isolation of naïve T cells. T cells were labeled with 10 uM CFDA (Invitrogen, Waltham, MA) (15 min, 37°C, 5% CO_2_), washed with media and incubated (30 min, 37°C, 5% CO_2_). Tregs and naive T cells were plated at a density of 5×10^4^ cells/well on a 96-well plate pre-coated with 5 ug/mL anti-CD3 (BD Pharmingen 145-2C11, Franklin Lakes, NJ) (2 hours, 37°C, 5% CO_2_). APCs were isolated using the same technique as above, but solely with PE anti-CD3 (BD Pharmingen 145-2C11, Franklin Lakes, NJ), anti-PE microbeads, and LD column. Isolated APCs were then incubated with 10 uL of 1 mg/mL Mitocycin C (ThermoFisher, Waltham, MA) for every 1×10^7^ cells (20 min, 37°C, 5%CO_2_). The APCs were then washed thrice with media and plated at a density of 1×10^5^ cells per well. Where indicated, 0.2 ng/mL IL-9 (Biolegend, San Diego, CA), 10 ng/mL anti-IL-9 (BE0181, BioXCell, Lebanon, NH), or 10 ng/mL isotype control (BE0085, BioXCell, Lebanon, NH) were added to the wells. After a 3 day incubation (37°C, 5% CO_2_), cells were incubated with 50 ng/mL PMA (Sigma-Aldrich, St. Louis, MO), 500 ng/mL Ionomycin (Sigma-Aldrich, St. Louis, MO), and Golgi Stop (BD Biosciences Franklin Lakes, NJ, Franklin Lakes, NJ) (3, hours, 37°C, 5% CO_2_). Cells were blocked with FC Block (Biolegend, San Diego, CA) per manufacturer’s instructions then stained for flow cytometry for the following: BV605 anti-CD3e (Biolegend 145-2C11, San Diego, CA), PE-Cy7 anti-CD4 (Biolegend RM4-5, San Diego, CA), APC anti-IFN_γ_ (Biolegend XMG1.2, San Diego, CA), PerCP-Cy5.5 anti-IL-17 (Biolegend TC11-18H10.1, San Diego, CA), AF488 anti-IL-4 (Biolegend 11B11,San Diego, CA), and BV421 anti-IL-9 (Biolegend RM9A4, San Diego, CA). Flow cytometry was performed on BD Fortessa with analysis on FlowJo X.(FlowJo, Waltham, MA).

### Th9 and Th17 polarization

To polarize T cells, B6 and B6.NOD-*Aec1Aec2* splenocytes were processed as above. Naive T cells were added at a density of 5×10^^4^ cells/well on a 96-well plate pre-coated with 5 μg/mL anti-CD3 (BD Pharmingen 145-2C11, Franklin Lakes, NJ) (2 hours, 37°C, 5% CO_2_). To all wells was added 2.5 ug/mL anti-CD28 (BD Pharmingen 37.51, Franklin Lakes, NJ). For Th9 polarization, 3 ng/mL TGFb1 (Biolegend, San Diego, CA) and 10 ng/mL IL-4 (Biolegend, San Diego, CA) were added to the wells; Th17 polarization, 3 ng/mL TGFb1 (Biolegend, San Diego, CA) and 30 ng/mL IL-6 (Biolegend, San Diego, CA) were added to the wells. After a 3-day incubation (37°C, 5% CO_2_), cells were incubated with 50 ng/mL PMA (Sigma-Aldrich, St. Louis, MO), 500 ng/mL Ionomycin (Sigma-Aldrich, St. Louis, MO), and Golgi Stop (BD Biosciences Franklin Lakes, NJ, Franklin Lakes, NJ) (3, hours, 37°C, 5% CO_2_). After the cells were blocked with FC Block (Biolegend, San Diego, CA) per manufacturer’s instructions, staining for flow cytometry was performed for the following: BV605 anti-CD3e (Biolegend 145-2C11, San Diego, CA), PE-Cy7 anti-CD4 (Biolegend RM4-5, San Diego, CA), APC anti-IFN_γ_ (Biolegend XMG1.2, San Diego, CA), PerCP-Cy5.5 anti-IL-17 (Biolegend TC11-18H10.1, San Diego, CA), AF488 anti-IL-4 (Biolegend 11B11,San Diego, CA), and BV421 anti-IL-9 (Biolegend RM9A4, San Diego, CA).Flow cytometry was performed on BD Fortessa with analysis on FlowJo X.(FlowJo, Waltham, MA).

### Statistical Analysis

Statistical analyses were performed using one-way ANOVA and Mann-Whitney test where indicated (Prism 8, GraphPad, La Jolla, CA). P valuesL<L0.05 were considered significant.

## RESULTS

### Elevated serum IL-9 levels and clinical correlations in SjD patients

IL-9 plays a crucial role in the immune system, particularly in the development and function of T helper cells. It is primarily produced by Th9 cells, and elevated levels of IL-9 have been observed in various autoimmune diseases, including RA, MS, and SLE. In this study, we aimed to investigate the levels of IL-9 and its associated cytokines in patients with SjD. We collected serum samples from healthy controls and SjD patients (**Supplemental Table 1**) and used a Bio-Plex multiplex immunoassay to measure cytokine levels. As shown in **Fig. 1**, IL-9 levels were significantly elevated (more than 10-fold) in SjD patients compared to healthy controls. We then examined the levels of IL-9/Th9-associated cytokines and found increased levels of IL-1α, IL-5, IL-6, IL-13, and TNF-α, with no significant differences in IL-10 levels between healthy controls and SjD patients.

**Figure 1:**
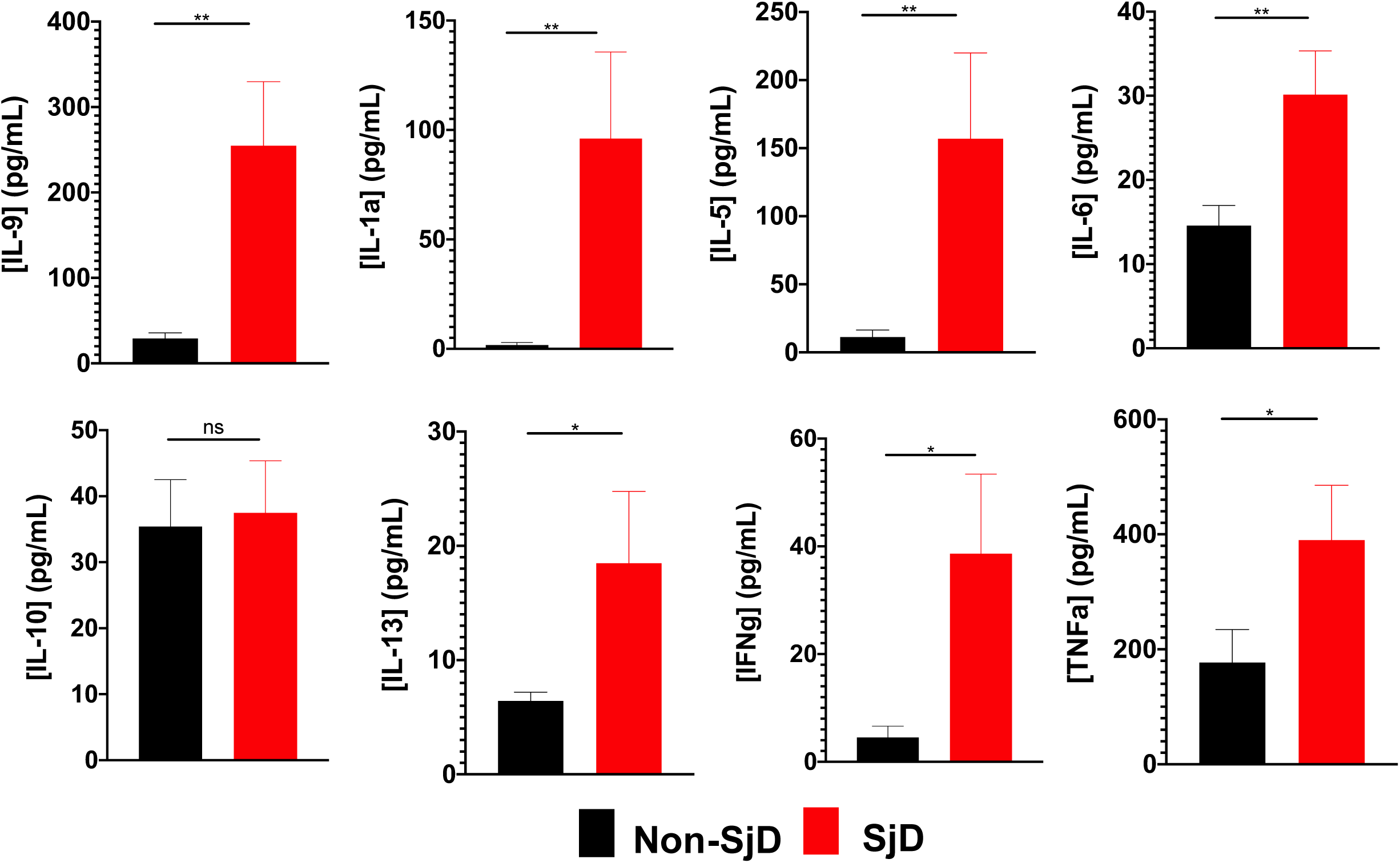
Elevation of serum cytokine levels in SjD patients. Cytokine analysis was performed on SjD patients (red) and controls (black), n=15 each group. Significance was determined by Mann-Whitney t-tests, where *p < 0.05 and **p < 0.01.

To evaluate the relationship between IL-9 and clinical laboratory results, we analyzed the correlation between IL-9 serum levels and various serological and diagnostic parameters. Our results showed that increased IL-9 levels were strongly correlated with several clinical laboratory parameters, including IgG, RF, ESR, ANA, ESSDAI, Ro52, Ro60, La, C3, C4, IgA, and IgM (**Table 1**). Notably, we observed a negative correlation between IL-9 levels and ESR, C3, C4, and IgA, where these values decreased as IL-9 levels increased. In contrast, a positive correlation existed between IL-9 levels and ANA, Ro60, IgM, and RF (**Fig. 2**). Overall, our findings indicate that SjD patients exhibit elevated levels of serum IL-9 and its associated cytokines, and there is a significant association between IL-9 levels and clinical laboratory parameters.

**Figure 2:**
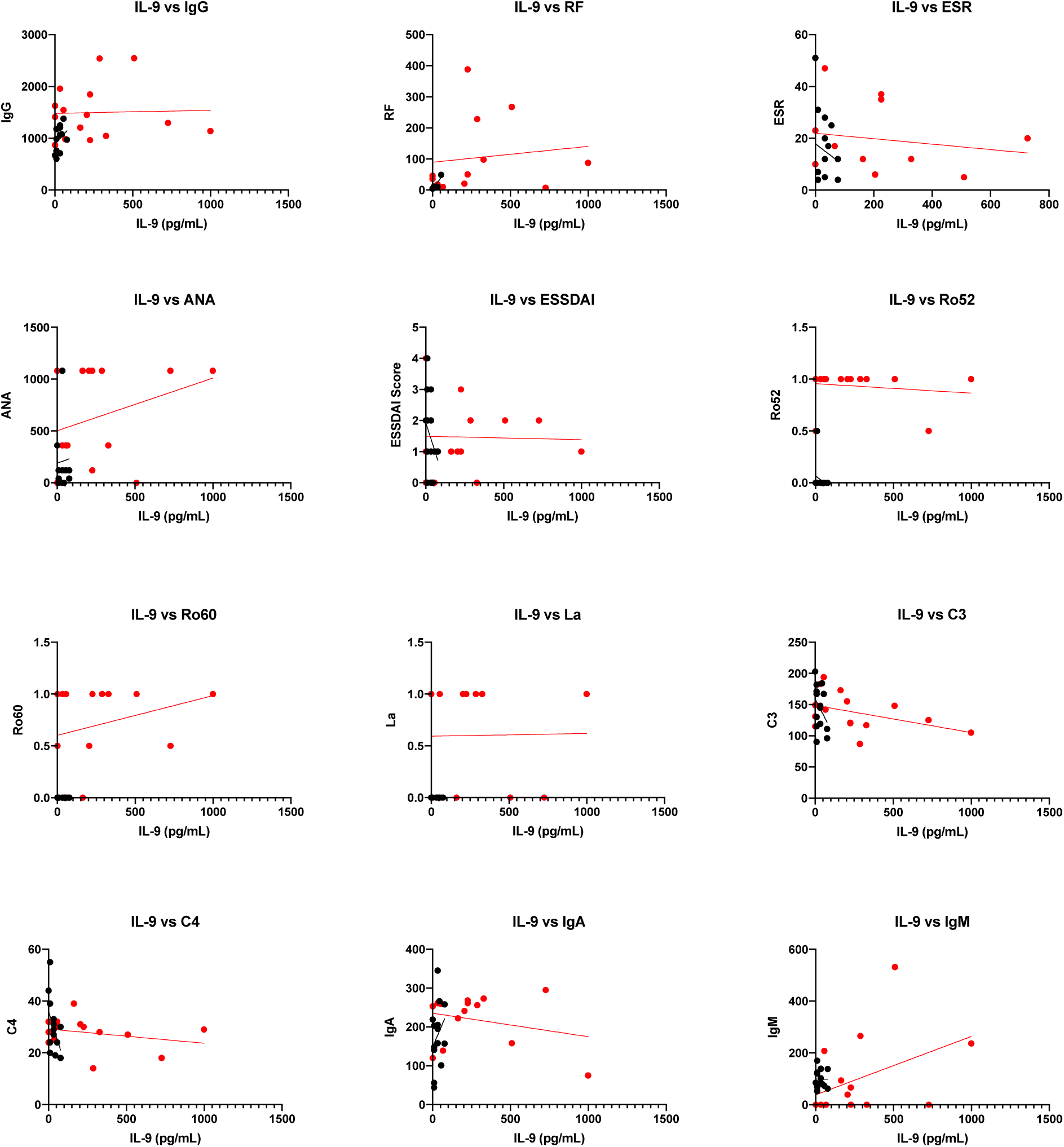
Correlation analysis of elevated IL-9 and patient clinical manifestations. The red line indicates the relationship between IL-9 levels of SjD (red) and control (black) patients in terms of clinical disease signs. A line sloping to the left indicates a negative correlation, a line sloping to the right indicates a positive correlation, and a line with no slope indicates no correlation. The significance of the correlation is denoted as *p < 0.05 and **p < 0.01.

**Table 1:** Clinical Association Between IL-9 and Clinical Laboratory Parameters.

| Patients | Spearman r | Significance | p-value |
| --- | --- | --- | --- |
| IgG | 1 | **** | <0.0001 |
| RF | 1 | **** | <0.0001 |
| ESR | 1 | **** | <0.0001 |
| ANA | 1 | **** | <0.0001 |
| ESSDAI | 1 | **** | <0.0001 |
| Ro52 | 1 | ** | 0.0095 |
| Ro60 | 1 | **** | <0.0001 |
| La | 1 | *** | 0.0002 |
| C3 | 1 | **** | <0.0001 |
| C4 | 1 | **** | <0.0001 |
| IgA | 1 | **** | <0.0001 |
| IgM | 1 | **** | <0.0001 |

### Elevated levels of IL-9 and Th9-associated cytokines in SjD mice

Studies suggest that IL-9 may worsen disease symptoms by promoting inflammation and supporting the activity of autoreactive T cells. Notably, IL-9 can enhance the pathogenic functions of Th9 cells, thereby amplifying the autoimmune response. Despite this, IL-9 levels have not been previously investigated in animal models of SjD. We collected sera from young (4-6 weeks of age) and old (24-30 weeks of age) control B6 and SjD mice. We conducted immunoassays to measure IL-9 levels and the associated cytokines produced by Th9 cells in SjD mice. Our results showed that serum IL-9 was significantly elevated in older SjD mice compared to healthy older control mice (B6 mice), with levels reaching 1267 ± 409.9 versus 476.6 ± 44.96 (**Fig. 3A**). Furthermore, we examined the levels of Th9-associated cytokines, including IL-1α, IL-4, IL-5, IL-6, IL-10, and GM-CSF. As shown in **Fig. 3B**, IL-1α, IL-4, IL-6, IL-10, and GM-CSF levels were significantly higher in older or diseased SjD mice, while IL-5 levels were also elevated compared to age– and sex-matched B6 mice. These findings indicate that IL-9 and Th9-associated cytokines are elevated in SjD mice.

**Figure 3:**
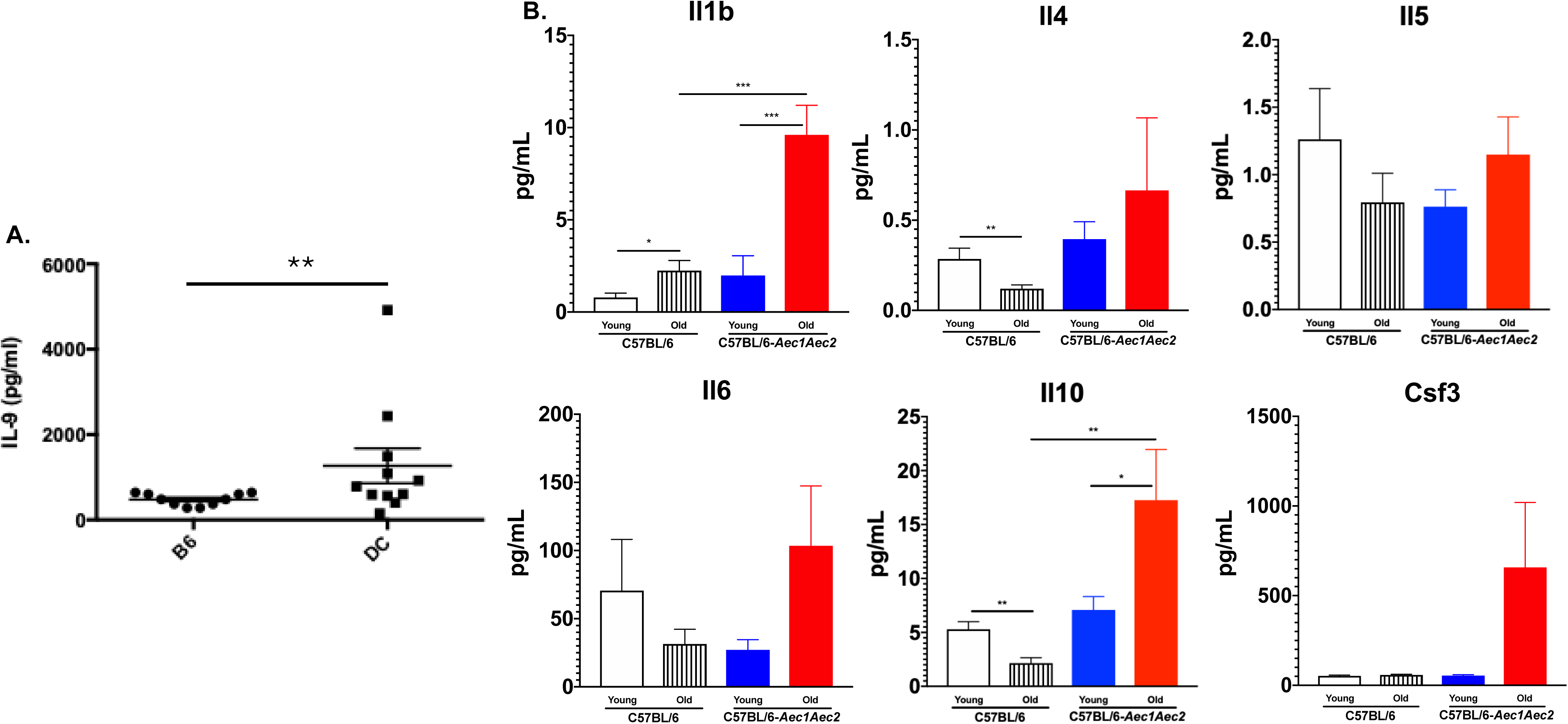
Elevation of serum cytokines in SjD mouse. Cytokine analysis was performed on SjD (square, n=11) and control (circle, n=10) mice for IL-9 (A). Further analysis was performed on young (white, n=5) and old (striped, n=5) control mice versus young (blue, n=5) and old (red, n=5) SjD mice for the indicated cytokines. Significance was determined by Mann-Whitney t-tests were performed for IL-9, and one-way ANOVA was used for other cytokines, where *p <0.05 and **p <0.01.

### Macrophages, NK cells, and T cells are responsible for IL-9 production

In addition to Th9 cells, IL-9 is produced by CD4^+^ T cells, CD8^+^ T cells, B cells, NKT cells, innate lymphoid cells (ILCs), and mast cells. As presented, IL-9 levels were significantly increased in SjD patients and mice. Therefore, we sought to determine the cellular source of IL-9 secretion in the SjD and compared it to the control B6 mice. We isolated immune cells from blood, cervical draining lymph nodes, salivary glands, and spleens of B6 and SjD mice. Flow cytometric and single-cell analyses were conducted to determine the immune cells positive for IL-9. As presented in **Fig. 4**, salivary glands appear to be the dominant source of IL-9^+^ cells that were expressed by CD11^+^ macrophage, CD19^+^ B cells, CD4^+^ T cells, and NKp46^+^ NK cells in comparison to blood, cervical LN, and spleen. Two notable observations in which B6 showed an elevated frequency of IL-9^+^ cells compared to SjD mice, and salivary gland effector Th17 cells showed the highest frequency of IL-9^+^ cells (approximately 60% in B6 and 40% in SjD mice). To further elucidate this relationship, single-cell analysis was performed on the salivary glands of B6 and SjD mice (**Fig. 5**). Reanalysis of GSE26853^10^ identified seven distinct major cell types, including macrophages, B cells, T cells, and NK cells. Lower Cd3e expression separated innate lymphoid cells from the major T cell groups, and Tcrg-C1 was used to identify gamma delta T cells (**Fig. 5A**). We examined the expression of transcription factors associated with Th9 cells across different cell types (**Fig. 5B**). Irf4 was highly expressed in B cells, while Irf8 was highly expressed in B cells, NK cells, and macrophages. Stat5a was expressed predominantly in T cell populations, similar to Ifng and Batf. Il17a was highly and exclusively expressed in gamma delta T cell populations, while Pparg was exclusively expressed in macrophages. Since Spi1 expression was primarily observed in macrophages and B cells (**Fig. 5C**), we compared its expression between B6 and SjD mice at the cell type level. Expression levels were comparable between macrophages of both strains, while B cells showed slightly higher Spi1 expression in SjD mice compared to B6 mice (percentage of expression: 26.7% vs. 18.9%, **Table 2**). UCell scores were calculated for individual cells based on Th9 transcription factor signatures using Mann-Whitney U statistical analysis (**Fig. 5D**). Single-cell level module analysis revealed that Th9 transcription factors were enriched in most cell groups except CD8^+^ T cells and innate lymphoid cells. B cells and CD4^+^ T cells showed higher Th9 scores in SjD mice, while NK cells demonstrated lower scores.

**Figure 4:**
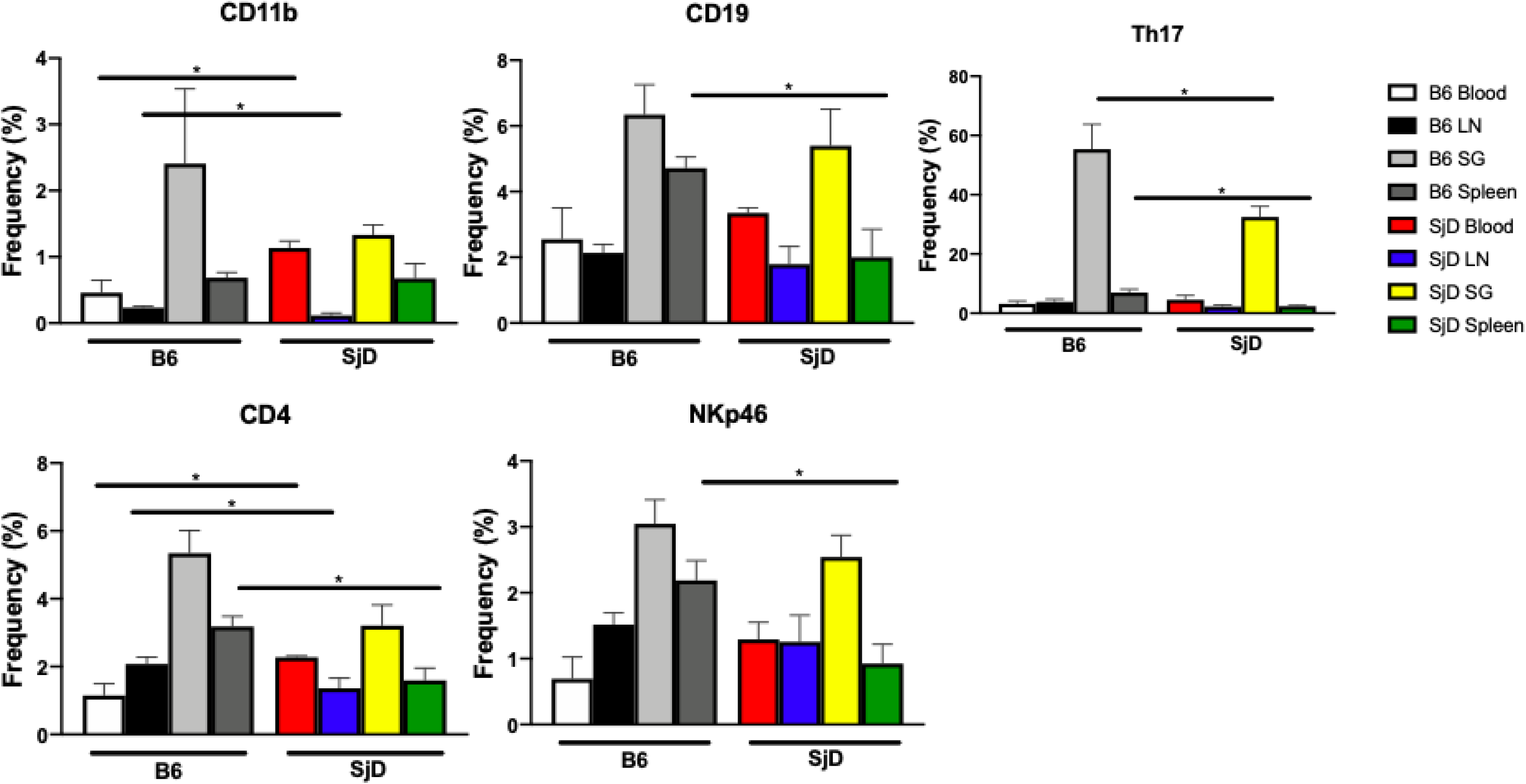
Cellular sources of IL-9 cytokines in SjD mouse by flow cytometry. Flow cytometry was utilized to determine cellular sources of IL-9 in the blood (white), lymph nodes (black), salivary glands (light gray), and spleen (dark gray) in control mice, as well as the corresponding sources in the SjD mice (blood (red), lymph nodes (blue), salivary glands (yellow), and spleen (green)), n= 3 mice/group. Mann-Whitney t-tests were performed, where *p <0.05.

**Figure 5.**
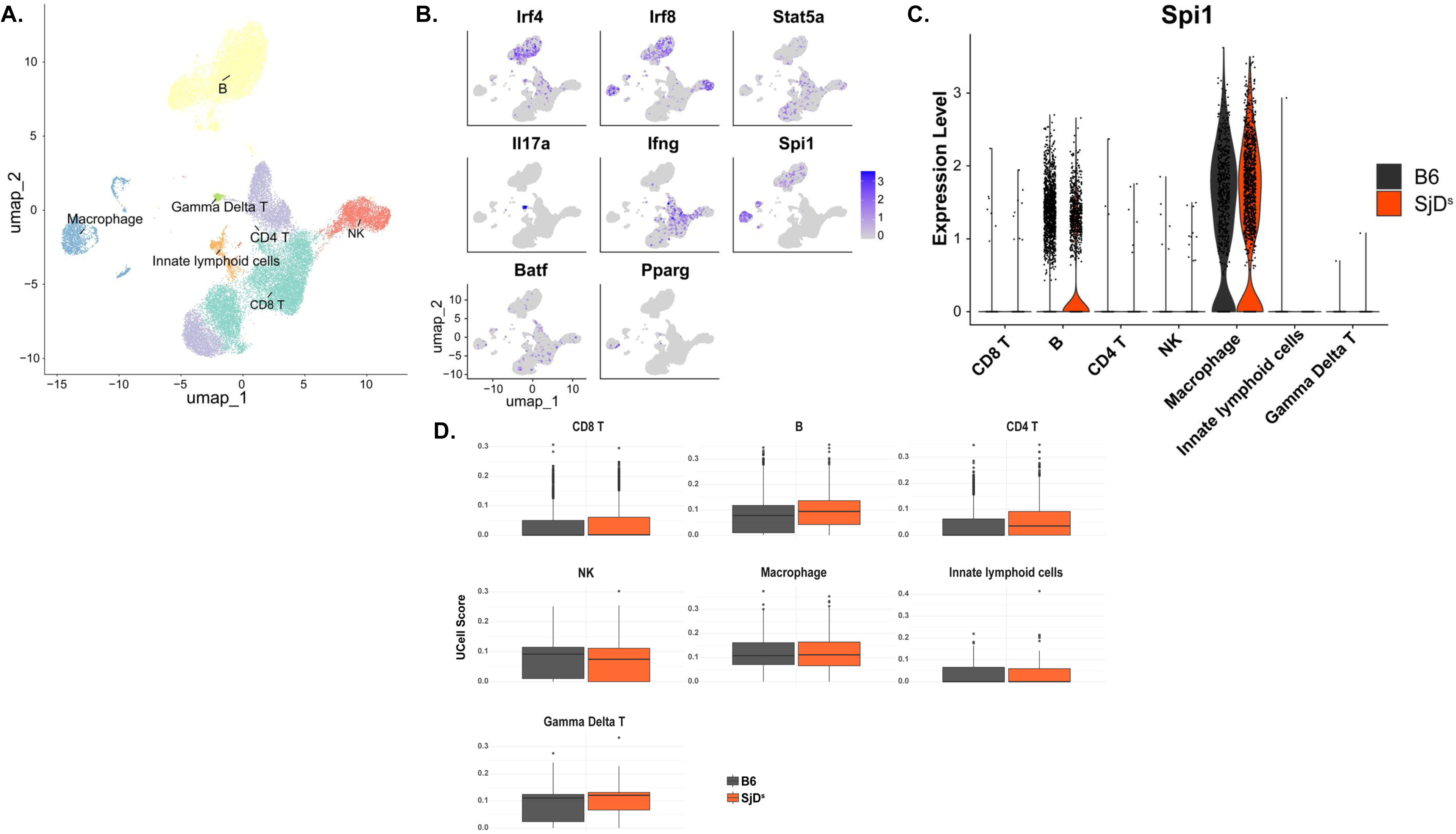
Cellular sources and transcriptional signatures of Th9-related genes in control and SjD-susceptible mice. (A) UMAP (Uniform Manifold Approximation and Projection) plots of immune cells showing seven major cell types identified in eight mice (B6 control mice, n = 2 of each sex; SjD^S^ (Susceptible) B6.NOD-*Aec1*/*2* mice, n = 2 of each sex). Different colors represent distinct cell populations. (B) Feature plots displaying the expression levels of Th9-associated transcription factors across different cell clusters. Color intensity represents the level of gene expression within each cluster. (C) Violin plots comparing Spi1 expression between SjD-susceptible mice and control mice across cell types, highlighting the primary cellular sources of Spi1 expression. (D) UCell scores for Th9 transcriptional signatures calculated using genes from the Th9 transcription factor network, showing cell type-specific enrichment patterns between mouse strains.

**Table 2:** Percentage of Sp1 cell type phenotype.

| **Strains** | **B cells** | **CD4<sup>+</sup> T** | **CD8<sup>+</sup> T** | **$\gamma\delta$ T** | **ILC** | **Macrophage** | **NK** |
| --- | --- | --- | --- | --- | --- | --- | --- |
| **B6** | 18.95 | 0.08 | 0.11 | 1.00 | 0.24 | 76.63 | 0.68 |
| **SjD** | 26.73 | 0.23 | 0.17 | 0.66 | 0 | 73.14 | 0.58 |

### Th9 and Th17 polarizations promote Th2, Th9, and Th17 cells in SjD mice

Th9 and Th17 shared several features, specifically the ability to secrete IL-9 and TGF-β is required for their in vitro differentiation^11,12^. A study has shown that Th17 cells from type 1 diabetic patients produced more IL-9, with a significant increase in IL-17^+^IL-9^+^ phenotype in cultures of cells isolated from the diabetic subjects compared to healthy individuals^13^. To determine whether IL-9– and IL-17-producing CD4 cells were altered in an autoimmune condition, we examined mice with SjD. Naïve T cells were isolated from healthy B6 mice and SjD mice. Cells stimulated with anti-CD3/CD28, TGF-β, and IL-4 for Th9 differentiation and anti-CD3/CD28, TGF-β, and IL-6 for Th17 differentiation. Naïve T cells of SjD mice showed a significant increase in Th1 under anti-CD3/28 and Th17-polarizing conditions (**Fig. 6A**). Th9-polarizing conditions drastically increased Th2 cells in SjD mice compared to healthy B6 mice (**Fig. 6B**). Both Th9– and Th17-polarizing conditions were able to stimulate the differentiation of Th9 cells in SjD mice (**Fig. 6C**). Interestingly, the Th9-polarizing condition showed a stronger impact on Th17 differentiation than the Th17-polarizing condition alone (**Fig. 6D**). Overall, our data indicate that SjD mice exhibit an increased propensity for Th1. And Th9-polarizing stimulation promotes Th2 and Th17, while a Th17-polarizing condition increases Th9 cells.

**Figure 6:**
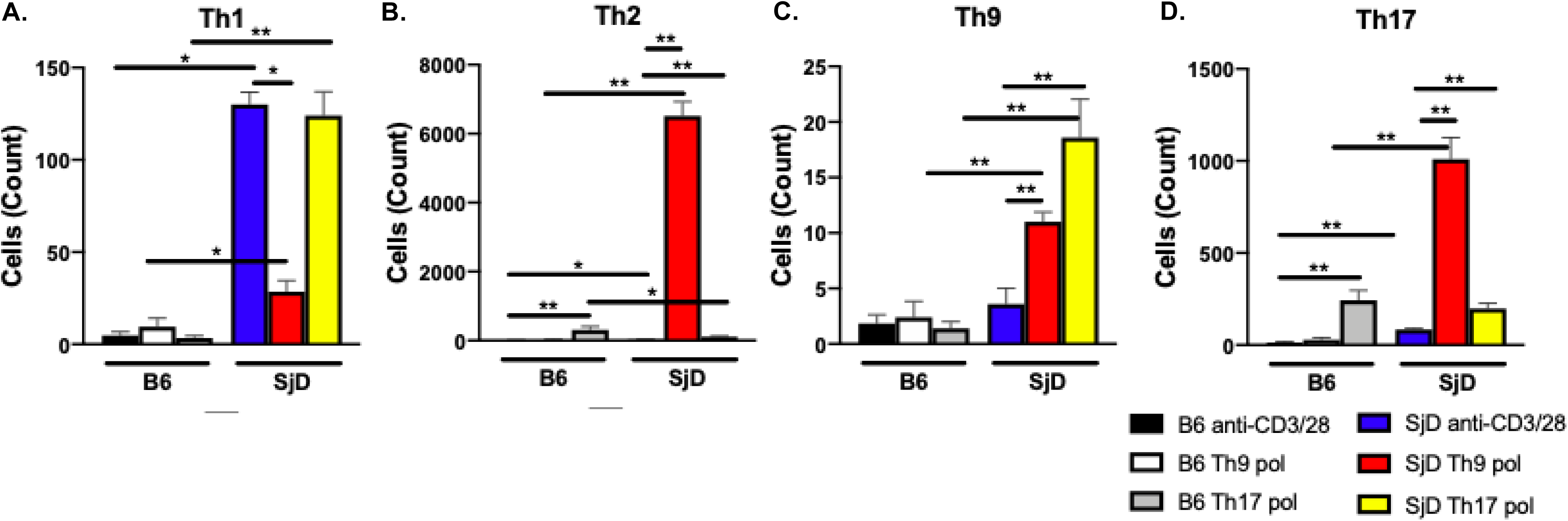
Ability of SjD mice to produce T helper cells under Th9 and Th17 polarization conditions. Polarization of naive T cells under non-antigenic or Th9 Th17 polarization conditions in both control (black, white, and gray, respectively) or SjD (blue, red, and yellow, respectively) mice. Resulting T helper populations are indicated (A Th1, B Th2, C Th9, and D Th17) with n=5/group. Mann-Whitney t-tests were performed, where *p <0.05 and **p <0.01.

### The effect of anti-IL-9 treatment on the disease signs of SjD mice

Our data have so far suggested that IL-9 and Th9 cells were elevated in SjD patients and mice. Th9 polarization promotes pathogenic Th2 and Th17. Moreover, studies have shown that IL-9 monoclonal antibody neutralization ameliorates CIA in DBA/1J mice^14^ and EAE in the MOG_35-55_ immunization model^15^. Therefore, we sought to determine the clinical effect of treating SjD with an anti-IL9 antibody. The SjD mice, B6.NOD-*Aec1Aec2*, received an IP injection of either an anti-IL-9 antibody or an isotype antibody control five times per week for six weeks. During the six-week treatment, mice were monitored for saliva flow, and focal scores and ANA were determined at the endpoint. As presented in **Fig. 7A**, there were no significant changes in SFR in anti-IL-9-treated or control mice. When the salivary glands were examined for focal scores, the anti-IL-9 treatment group was significantly worse, with average focal scores almost doubling compared to the control group (**Fig. 7B**). However, examination of the ANA staining patterns revealed a moderate improvement with a shift from only 13% negative to 36% negative with anti-IL-9 (**Fig. 7C**). To further evaluate if the mixed response to the anti-IL-9 treatment was due to the previously discovered sexual dimorphism in the SjD mice, we analyzed the mice separated by their sexes. There was only a slight improvement in SFR at week 2 between the females, with the treatment group showing a slight increase above the isotype control (**Fig. 7D**). However, no discernible difference was observed between the SFRs, either between the sexes or in response to the treatment, at 4 weeks post-treatment. The focus scores were higher in females treated with the anti-IL-9 antibody (**Fig. 7E**). Notably, while the other groups had an average at or below 2, the anti-IL-9-treated female group had an average of 6, indicating an influx of lymphocytes into the salivary glands of these mice. In terms of the ANA, which had previously improved, analysis by sex revealed that this improvement was solely within the male anti-IL-9-treated group (**Fig. 7F**), where autoantibodies were detected in only half of the mice, compared to 86% in the male isotype controls. Female mice were unaffected in their autoantibody profile, with both isotype and anti-IL-9 treatments maintaining only 20% negative. These data indicate a sexually dimorphic mechanism of IL-9, where blocking this pathway results in an improvement of ANA in male mice, whereas this causes exacerbation of the SjD signs, specifically the focus score in female mice.

**Figure 7:**
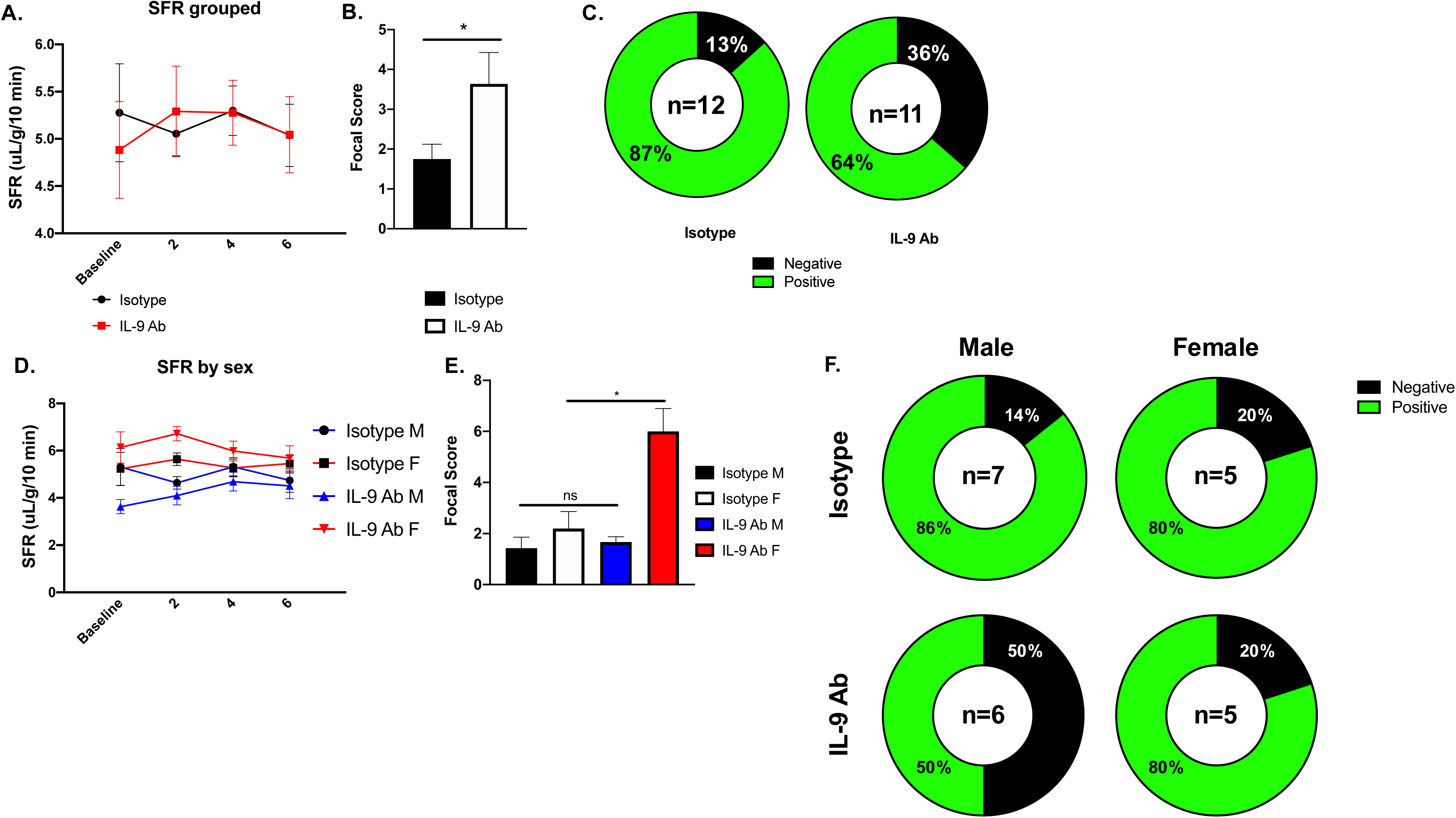
Treatment of SjD mice with anti-IL-9 does not improve phenotype. Either an isotype control or an anti-IL-9 antibody was administered to SjD mice, and diagnostic criteria were monitored. A. Saliva flow rate is indicated for the isotype (black), n=11, or anti-IL-9 antibody (i.e. IL-9 Ab, red), n=12, treatments. B. Focal scores are provided for the isotype (black) or IL-9 Ab (white). C. Anti-nuclear antibodies were evaluated in terms of negative (black) or positive (green) for the treatment indicated; percentage breakdown of each is provided. Further analysis is provided to elucidate if there was a sexually dimorphic response to the treatments. D. Saliva flow rate is indicated for the male (black circle with blue line, n= 7) or female (black square with red line, n= 5) isotype or IL-9 Ab male (blue circle with blue line, n= 6) or female (red square with red line, n= 6) treatments. E. Focal scores are provided for the isotype males (black) or females (white) or IL-9 Ab males (blue) or females (red). C. Anti-nuclear antibodies were evaluated in terms of negative (black) or positive (green) for the treatment indicated; percentage breakdown of each is provided.

### Tregs failed to suppress Th17 cells when IL-9 was blocked

A study has shown that IL-9 induces differentiation of Th17 cells and enhances the function of natural Tregs. As demonstrated, blocking IL-9 exacerbated the SjD phenotype; therefore, we sought to determine if Tregs’ suppressive function was being disrupted by treating with anti-IL-9. Here, a Treg suppression assay was performed to understand if the functional ability of Tregs is inherently different in the SjD mice compared to control mice and if the addition or blocking of IL-9 can affect this activity. When measuring the overall proliferation of T cells, Tregs from SjD mice behaved similarly to control mice. Addition of IL-9 trended toward less proliferation in both SjD and B6 mice. Interestingly, adding anti-IL-9 appeared to block Treg ability to suppress T cells, resulting in significant proliferation (**Fig. 8A)**.

**Figure 8:**
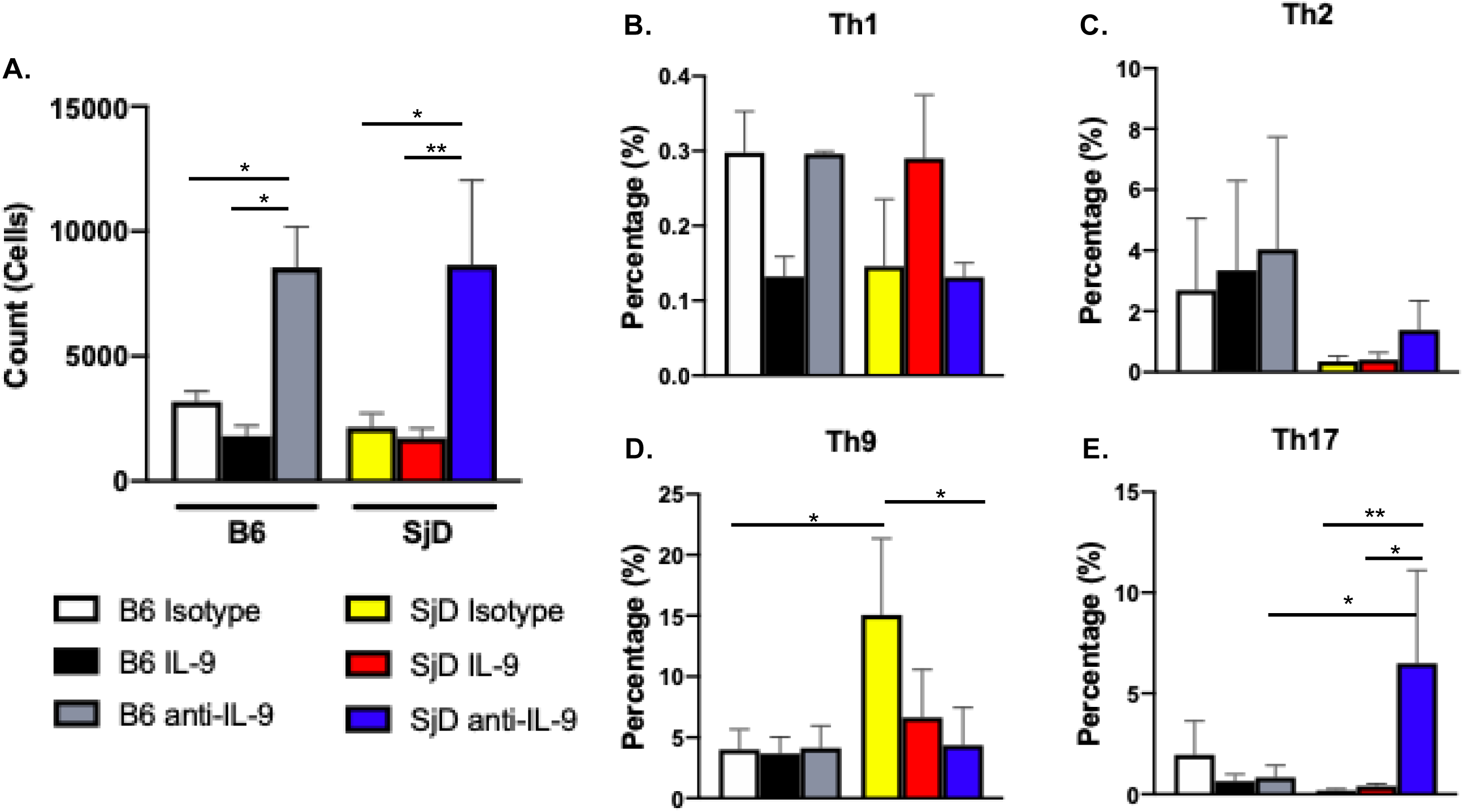
Tregs fail to suppress Th17 cells in SjD mice when IL-9 is blocked. Evaluation of whether Tregs suppressive function is enhanced by IL-9 or inhibited by anti-IL-9 compared to the isotype control. A. Total cells produced in control or SjD mice in the presence of isotype control (white or yellow, respectively), IL-9 (black or red, respectively), or anti-IL-9 (gray or blue, respectively). B-D The percentage of these cells broken down by T helper subset (Th1, Th2, Th9, or Th17) in the presence of the aforementioned conditions in both mouse lines, n=5/group. Kruskal-Wallis was performed for significance, where *p <0.05 and **p <0.01.

To determine if there were specific subsets of T effector cells affected by Tregs in response to IL-9 conditions, or if it is a systemic effect in all effector T cells. As expected in the healthy control B6 mice, Tregs were able to suppress Th1 proliferation when treated with IL-9, whereas Th1 proliferation was accelerated when treated with anti-IL-9. In the SjD mice, the results seemed to be the opposite, in which adding IL-9 increased Th1 proliferation, and adding anti-IL-9 decreased Th1 proliferation (**Fig. 8B**). Th2 cells were not affected by IL-9 or anti-IL-9 treatments in both B6 and SjD mice (**Fig. 8C),** whereas Th9 cells were negatively affected by both treatments (**Fig. 8D**). Interestingly, Th17 cells of SjD mice were significantly increased in proliferation when treated with anti-IL-9 in comparison to the isotype control, IL-9 treatment, or B6 mice with similar treatments (**Fig. 8E**). The data suggest that there are some intrinsic abilities of how Tregs respond to IL-9 to perform their suppressive function in SjD mice and blocking IL-9 may compromise Treg function.

## DISCUSSION

This study aimed to investigate the role of IL-9 and the Th9 axis in SjD across human subjects and a well-characterized mouse model, and to elucidate the functional consequences of IL-9 in the autoimmune model. We have found that, first, serum IL-9 is markedly elevated in human SjD and associates with a broad constellation of serological features central to disease activity. Second, IL-9 and Th9-associated cytokines are similarly elevated in SjD mice, supporting the notion that IL-9 dysregulation is a common feature in both humans and mice with SjD. Third, the cellular source of IL-9 in the SjD context appears to be more diverse than a canonical Th9 narrative would suggest, with macrophages, CD4^+^ T cells, and NK cells contributing in peripheral blood and a myeloid transcriptional imprint (Spi1/PU.1). Fourth, T helper cell plasticity in SjD is striking, in which Th9– and Th17-polarizing milieus do not simply yield their canonical linages, but instead boost Th2, Th9, and Th17 subsets in a manner that is heightened in SjD over controls. Finally, anti-IL-9 neutralization produces a subtle and sex-dependent pattern of effects in vivo, decreasing ANA positivity in males while worsening focal glandular infiltration in females, and impairing Treg-mediated control of Th17 proliferation ex vivo.

The more than tenfold elevation of serum IL-9 in SjD patients underscores IL-9 as a potentially important cytokine in SjD. Moreover, IL-9 correlates with ANA, Ro60, IgM, and RF, supporting an association between IL-9 activity and the B cell/autoantibody in SjD. IL-9 is known to enhance B cell responses, including promoting the exit of precursor memory B cells (MBC) from germinal centers and development into MBCs^16^. It promotes B-cell proliferation and differentiation from naïve B cells to plasma cells^17^. Recent data indicate that it is involved in class switching to IgE and IgG responses^18^. A positive correlation between IL-9, IL-4, and IL-6 (all of which are elevated in the mouse model) further supports their function in B cell activation and autoantibody production. The IgA negative correlation may align with IL-9 skewing class-switch recombination away from IgA toward IgG response in an IL-4-rich environment, or with mucosal glandular dysfunction limiting IgA production and translocation despite systemic inflammation^19^. C3 and C4 consumption is a classic consequence of immune complex formation. If IL-9 promotes autoantibody production, immune complexes could rise and drive complement consumption, explaining the inverse relationship between IL-9 and C3/C^20,21^. The SjD animal model replicated the patient observation, in which systemic IL-9 is elevated, and the increase co-occurred with higher levels of IL-1α, IL-4, IL-6, IL-10, GM-CSF, and IL-5—particularly in older and disease-manifested animals. IL-1α and IL-6 are potent drivers of Th17 differentiation and myeloid activation^22,23^. GM-CSF stabilizes inflammatory circuits between T cells and myeloid cells, often promoting tissue-destructive macrophage phenotypes^24^. IL-4 and IL-5 signal Th2 activity, which can support B-cell help and the type of tissue remodeling sometimes seen in chronically inflamed glands^25^. The co-elevation of these cytokines with IL-9 suggests a tightly connected network: IL-9 may not act alone but rather reinforce an inflammatory milieu in which myeloid cells and helper T cells co-amplify each other’s effector programs.

The observation that Th17 cells producing IL-9 are more frequent in the salivary glands of control mice and SjD-susceptible mice, and are higher in control mice, is unique. It may indicate that in healthy tissue, a subset of Th17 cells co-producing IL-9 serves a regulatory or homeostatic role^26^, whereas in disease, this subset contracts and is replaced by other IL-9 sources that skew pathogenic^27^. Alternatively, SjD may drive Th17 cells toward a more classical IL-17-only phenotype while shifting IL-9 production to myeloid or circulating T cell pools. Single-cell transcriptomics indicate the expression of Th9-associated transcription factors by immune cells found in the salivary gland, specifically, the critical Spi1 expression was comparable in both normal and SjD mice. However, using UCell analysis, which accounted for all published Th9-related transcription factors, B cells, and CD4^+^ T cells, showed higher Th9 scores in SjD mice. These two cell populations are the major drivers of autoimmune response in SjD, in terms of autoantibody production and effector T cell function. Recent data have demonstrated that IL-9 is required for humoral memory recall due to its promoting effect on the development and function of memory B cells from germinal centers. Additionally, T cell-derived IL-9 is required for the education of functionally competent memory B cells in the primary response^28^.

The in vitro polarization data showed a remarkable plasticity of CD4^+^ T cells in SjD. Exposure of naïve T cells from SjD mice to Th9-polarizing conditions not only yielded more Th9 cells, but it also drove a substantial rise in Th2 cells and enhanced Th17 differentiation more strongly than canonical Th17-polarizing conditions did. Conversely, Th17-polarizing conditions increased Th9 cells in SjD. These reciprocal effects suggest a shared epigenetic program in SjD T cells that renders them permissive to multiple differentiation pathways along the Th2/Th9/Th17 axis when TGF-β is present^13,29,30^. Salivary glands express a high level of TGF-β, an important cytokine for gland development and immune cell modulation, and its level is upregulated in SjD salivary glands^31^. Therefore, the level and availability of TGF-β can dictate the type of immune response required, whether to maintain homeostasis, regulate, or induce pathogenicity. In an autoimmune setting like SjD, such plasticity may be modulated by chronic antigen exposure, defective inflammation resolution, and altered metabolic states, which could lower the thresholds for activation and cell lineage interconversion^32–34^. Functionally, this plasticity may help explain the observed cytokine milieu. A Th9-primed microenvironment may amplify both Th2 and Th17 pathology. Th2 responses contribute to B cell help and tissue remodeling, while Th17 responses recruit neutrophils and monocytes and can be particularly destructive in exocrine tissues. The increased propensity for Th1 differentiation in SjD further expands the inflammatory milieu, enabling IFN-γ to synergize with IL-6 and GM-CSF in activating macrophages.

The anti-IL-9 intervention experiments reveal a complex therapeutic approach to SjD. Despite high systemic IL-9, neutralization did not improve salivary flow rates, suggesting that glandular functional impairment in this model is not primarily limited by IL-9-dependent mechanisms, at least under the treatment regimen, mouse ages, and dosing tested. More strikingly, anti-IL-9 treatment increased focal lymphocytic infiltration in the salivary glands, nearly doubling the focus scores overall, and this effect was predominantly observed in females. Yet anti-IL-9 reduced ANA positivity in males. This dissociation, worsened focal inflammation alongside improved autoantibody status in males, implies that IL-9 simultaneously participates in local immunoregulation that restrains tissue infiltration and in systemic autoantibody response that sustains ANA production. Removing IL-9 can therefore shift the balance in opposite directions depending on the clinical signs measured and the sex of the mouse model. If IL-9 supports Treg function (as the ex vivo data suggest), females may rely more heavily on IL-9-dependent Treg pathways to restrain glandular Th17-driven inflammation. Blocking IL-9 would preferentially impair this restraint in females, increasing infiltrates. In males, relatively stronger androgenic support for Treg stability or reduced Th17 pressure may mitigate the loss of IL-9, allowing for a net benefit on autoantibody response without promoting local inflammation^35^. The reduction in ANA positivity in males suggests that in male mice, IL-9 fosters B cell autoimmunity more directly. If female B cell responses are less IL-9-dependent or if compensatory pathways dominate (for instance, BAFF-driven survival), IL-9 blockade might not reduce ANA in females, consistent with the data.

Studies have shown that IL-9 plays an integral function in enhancing regulatory T cell^26,30^. As expected, anti-IL-9 compromised Treg suppression of T cell proliferation, indicating that IL-9 contributes to the immunosuppressive capacity of Tregs. In subset T cell analyses, the most consistent and disease-relevant pattern was a loss of control over Th17 proliferation in SjD when IL-9 was blocked. Given the well-established pathogenicity of Th17 cells in SjD^36,37^, this provides a plausible causal link to the worsened salivary gland infiltration seen in females: blocking IL-9 undermines Treg restraint of Th17, thereby enhancing glandular inflammation, as observed with a higher focus score. Th2 proliferation was largely unaffected by IL-9 manipulations in both strains, while Th9 cells were negatively affected by both exogenous IL-9 and its blockade. One interpretation is that Tregs require IL-9 to maintain their suppressive potency against Th17 cells specifically; therefore, by stabilizing Foxp3 expression, they are relatively IL-9-independent, unlike Th2 cells. Taken together, these findings show IL-9 is an important cytokine in SjD pathobiology. Elevated in serum and linked to autoantibody production, IL-9 participates in a broader Th9/Th2/Th17 axis that is especially prominent with advancing disease in mice and human patients.

## AUTHOR CONTRIBUTIONS

PG maintained mouse lines, treated mice, and evaluated salivary flow rates. AV performed cytokine analyses, flow cytometry, antinuclear antibody assays, *ex vivo* Treg and naive T cell assays, and evaluated focal scores. YS conducted the single-cell analysis. AR, RHS, KG, CJL, and DF were responsible for patient recruitment, sample collection, and disease profiling. AV and CQN conceptualized, analyzed, and participated in the manuscript writing.

## Data Availability

All data produced in the present work are contained in the manuscript

## ACKNOWLEDGEMENTS

We would like to extend our gratitude to Lida Radfar and Donald U. Stone for their expertise in conducting the clinical evaluations, and to David Lewis for his valuable contributions to the pathological evaluations of the patients. Additionally, we appreciate the efforts of Kathy L. Sivils for establishing the Oklahoma Sjögren’s Research Clinic. This research was funded by the National Institutes of Health (NIH) and the National Institute of Dental and Craniofacial Research (NIDCR) (DE028544, DE028544-02S1, PI-Nguyen).

## REFERENCE

1 Ciccia, F. et al. Potential involvement of IL-9 and Th9 cells in the pathogenesis of rheumatoid arthritis. Rheumatology (Oxford) 54, 2264–2272, doi:10.1093/rheumatology/kev252 (2015).

2 Chowdhury, K. et al. Synovial IL-9 facilitates neutrophil survival, function and differentiation of Th17 cells in rheumatoid arthritis. Arthritis Research & Therapy 20, 18, doi:10.1186/s13075-017-1505-8 (2018).

3 Ouyang, H. et al. Increased interleukin⍰9 and CD4+IL-9+ T cells in patients with systemic lupus erythematosus. Mol Med Rep 7, 1031–1037, doi:10.3892/mmr.2013.1258 (2013).

4 Tu, J. et al. Positive feedback loop PU.1-IL9 in Th9 promotes rheumatoid arthritis development. Ann Rheum Dis 83, 1707–1721, doi:10.1136/ard-2024-226067 (2024).

5 Rasmussen, A. et al. Comparison of the American-European Consensus Group Sjögren’s syndrome classification criteria to newly proposed American College of Rheumatology criteria in a large, carefully characterised sicca cohort. Annals of the Rheumatic Diseases 73, 31–38, 10.1136/annrheumdis-2013-203845 (2014).

6 Vitali, C. et al. Classification criteria for Sjögren’s syndrome: a revised version of the European criteria proposed by the American-European Consensus Group. Ann Rheum Dis 61, 554–558, doi:10.1136/ard.61.6.554 (2002).

7 Shiboski, C. H. et al. 2016 American College of Rheumatology/European League Against Rheumatism Classification Criteria for Primary Sjögren’s Syndrome: A Consensus and Data-Driven Methodology Involving Three International Patient Cohorts. Arthritis Rheumatol 69, 35–45, doi:10.1002/art.39859 (2017).

8 Ulrich, B. J. et al. Allergic airway recall responses require IL-9 from resident memory CD4(+) T cells. Sci Immunol 7, eabg9296, doi:10.1126/sciimmunol.abg9296 (2022).

9 Kaplan, M. H. The transcription factor network in Th9 cells. Semin Immunopathol 39, 11–20, doi:10.1007/s00281-016-0600-2 (2017).

10 Shen, Y., Voigt, A., Bhattacharyya, I. & Nguyen, C. Q. Single-Cell Transcriptomics Reveals a Pivotal Role of DOCK2 in Sjögren Disease. ACR Open Rheumatol 6, 927–943, doi:10.1002/acr2.11738 (2024).

11 Manel, N., Unutmaz, D. & Littman, D. R. The differentiation of human T(H)-17 cells requires transforming growth factor-beta and induction of the nuclear receptor RORgammat. Nat Immunol 9, 641–649, doi:ni.1610 [pii] 10.1038/ni.1610 (2008).

12 Nowak, E. C. et al. IL-9 as a mediator of Th17-driven inflammatory disease. J Exp Med 206, 1653–1660, doi:10.1084/jem.20090246 (2009).

13 Beriou, G. et al. TGF-beta induces IL-9 production from human Th17 cells. J Immunol 185, 46–54, doi:10.4049/jimmunol.1000356 (2010).

14 Tu, J. et al. Positive feedback loop PU.1-IL9 in Th9 promotes rheumatoid arthritis development. Annals of the Rheumatic Diseases 83, 1707–1721, 10.1136/ard-2024-226067 (2024).

15 Nowak, E. C. et al. IL-9 as a mediator of Th17-driven inflammatory disease. Journal of Experimental Medicine 206, 1653–1660, doi:10.1084/jem.20090246 (2009).

16 Wang, Y. et al. Germinal-center development of memory B cells driven by IL-9 from follicular helper T cells. Nature immunology 18, 921–930 (2017).

17 Takatsuka, S. et al. IL-9 receptor signaling in memory B cells regulates humoral recall responses. Nature Immunology 19, 1025–1034, doi:10.1038/s41590-018-0177-0 (2018).

18 Sato, T. et al. Interleukin 9 mediates T follicular helper cell activation to promote antibody responses. Front Immunol 15, 1441407, doi:10.3389/fimmu.2024.1441407 (2024).

19 Nguyen, C. Q. et al. IL-4-STAT6 signal transduction-dependent induction of the clinical phase of Sjogren’s syndrome-like disease of the nonobese diabetic mouse. J Immunol 179, 382–390, doi:179/1/382 [pii] (2007).

20 Nguyen, C. et al. Role of complement and B lymphocytes in Sjogren’s syndrome-like autoimmune exocrinopathy of NOD.B10-H2b mice. Mol. Immunol. 43, 1332–1339 (2006).

21 Nguyen, C. Q., Kim, H., Cornelius, J. G. & Peck, A. B. Development of Sjogren’s syndrome in nonobese diabetic-derived autoimmune-prone C57BL/6.NOD-Aec1Aec2 mice is dependent on complement component-3. J Immunol 179, 2318–2329, doi:10.4049/jimmunol.179.4.2318 (2007).

22 Pyrillou, K., Burzynski, L. C. & Clarke, M. C. H. Alternative Pathways of IL-1 Activation, and Its Role in Health and Disease. Front Immunol 11, 613170, doi:10.3389/fimmu.2020.613170 (2020).

23 Huangfu, L., Li, R., Huang, Y. & Wang, S. The IL-17 family in diseases: from bench to bedside. Signal Transduction and Targeted Therapy 8, 402, doi:10.1038/s41392-023-01620-3 (2023).

24 Ingelfinger, F., De Feo, D. & Becher, B. GM-CSF: Master regulator of the T cell-phagocyte interface during inflammation. Seminars in Immunology 54, 101518, 10.1016/j.smim.2021.101518 (2021).

25 Gieseck, R. L., Wilson, M. S. & Wynn, T. A. Type 2 immunity in tissue repair and fibrosis. Nature Reviews Immunology 18, 62–76, doi:10.1038/nri.2017.90 (2018).

26 Elyaman, W. et al. IL-9 induces differentiation of TH17 cells and enhances function of FoxP3+ natural regulatory T cells. Proc Natl Acad Sci U S A 106, 12885–12890, doi:10.1073/pnas.0812530106 (2009).

27 Schlapbach, C. et al. Human TH9 cells are skin-tropic and have autocrine and paracrine proinflammatory capacity. Sci Transl Med 6, 219ra218, doi:10.1126/scitranslmed.3007828 (2014).

28 Luo, X. et al. An interleukin-9-ZBTB18 axis promotes germinal center development of memory B cells. Immunity 58, 861–874.e866, doi:10.1016/j.immuni.2025.02.021 (2025).

29 Korn, T., Bettelli, E., Oukka, M. & Kuchroo, V. K. IL-17 and Th17 Cells. Annu Rev Immunol 27, 485–517, doi:10.1146/annurev.immunol.021908.132710 (2009).

30 Chattopadhyay, S., Hazra, S., Biswas, B. & Goswami, R. Homeobox Transcription Factor HOXB4 Distinctly Modulates Th2 and Th9 Cell Differentiation. Eur J Immunol 55, e51752, doi:10.1002/eji.202451752 (2025).

31 Killedar, S. J. et al. Early pathogenic events associated with Sjogren’s syndrome (SjS)-like disease of the NOD mouse using microarray analysis. Laboratory investigation; a journal of technical methods and pathology 86, 1243–1260, doi:10.1038/labinvest.3700487 (2006).

32 Witas, R. et al. Defective Efferocytosis in a Murine Model of Sjögren’s Syndrome Is Mediated by Dysfunctional Mer Tyrosine Kinase Receptor. Int J Mol Sci 22, doi:10.3390/ijms22189711 (2021).

33 Chen, Y. et al. Immunometabolic alteration of CD4+ T cells in the pathogenesis of primary Sjögren’s syndrome. Clin Exp Med 24, 163 (2024).

34 Chen, B. et al. CD4+ T-cell metabolism in the pathogenesis of Sjogren’s syndrome. International Immunopharmacology 150, 114320, 10.1016/j.intimp.2025.114320 (2025).

35 Gandhi, V. D. et al. Androgen receptor signaling promotes Treg suppressive function during allergic airway inflammation. J Clin Invest 132, doi:10.1172/jci153397 (2022).

36 Nguyen, C. Q., Hu, M. H., Li, Y., Stewart, C. & Peck, A. B. Salivary gland tissue expression of interleukin-23 and interleukin-17 in Sjogren’s syndrome: Findings in humans and mice. Arthritis Rheum 58, 734–743, doi:10.1002/art.23214 (2008).

37 Nguyen, C. Q. et al. Pathogenic effect of interleukin-17A in induction of Sjogren’s syndrome-like disease using adenovirus-mediated gene transfer. Arthritis research & therapy 12, R220, doi:10.1186/ar3207 (2010).

