## Supplemental Table 1 for "Elevated Levels of IL-9 Fail to Suppress Pathogenic T helper 17 cells in Sjögren’s Disease"

**Table S1: Patient profiles for clinical samples.**

| Subjects | Age range (years) | Sex | Disease | Subjects | Age range (years) | Sex | Disease |
|---|---|---|---|---|---|---|---|
| 1 | 28-79 | Female | SjD | 1 | 30-78 | Female | Non-SjD |
| 2 | | Female | SjD | 2 | | Female | Non-SjD |
| 3 | | Female | SjD | 3 | | Female | Non-SjD |
| 4 | | Female | SjD | 4 | | Female | Non-SjD |
| 5 | | Male | SjD | 5 | | Male | Non-SjD |
| 6 | | Female | SjD | 6 | | Female | Non-SjD |
| 7 | | Female | SjD | 7 | | Female | Non-SjD |
| 8 | | Female | SjD | 8 | | Female | Non-SjD |
| 9 | | Female | SjD | 9 | | Female | Non-SjD |
| 10 | | Female | SjD | 10 | | Female | Non-SjD |
| 11 | | Female | SjD | 11 | | Female | Non-SjD |
| 12 | | Female | SjD | 12 | | Female | Non-SjD |
| 13 | | Female | SjD | 13 | | Female | Non-SjD |
| 14 | | Female | SjD | 14 | | Female | Non-SjD |
| 15 | | Female | SjD | 15 | | Female | Non-SjD |
